# A Phase 1b/2a Clinical Trial of Dantrolene Sodium in Patients with Wolfram Syndrome

**DOI:** 10.1101/2020.10.07.20208694

**Authors:** Damien Abreu, Stephen I Stone, Toni Pearson, Robert Bucelli, Ashley N Simpson, Stacy Hurst, Cris M Brown, Kelly Kries, Hongjie Gu, James Hoekel, Lawrence Tychsen, Gregory P. Van Stavern, Neil H White, Bess A Marshall, Tamara Hershey, Fumihiko Urano

## Abstract

**Background:** Wolfram syndrome is a rare endoplasmic reticulum disorder characterized by insulin-dependent diabetes mellitus, optic nerve atrophy, and progressive neurodegeneration. Although there is currently no treatment to delay, halt, or reverse the progression of Wolfram syndrome, preclinical studies in cell and rodent models suggest that therapeutic strategies targeting endoplasmic reticulum calcium homeostasis, including dantrolene sodium, may be beneficial.

**Methods:** Based on the results from preclinical studies on dantrolene sodium and ongoing longitudinal studies, our group put together the first-ever clinical trial in pediatric and adult patients with Wolfram syndrome. An open-label phase 1b/2a trial design was chosen. The primary objective of the study was to assess the safety and tolerability of dantrolene sodium in adult and pediatric patients with Wolfram syndrome. Secondary objectives were to evaluate the efficacy of dantrolene sodium on residual pancreatic β-cell functions, visual acuity, quality of life measures related to vision, and neurological functions.

**Results:** The results indicate that dantrolene sodium is well tolerated by patients with Wolfram syndrome. Although the study was small, a select few patients seemed to have improvements in β-cell function, which might correlate with a positive trend in other outcome measures, including visual acuity and neurological functions.

**Conclusion:** This study justifies further investigation into using dantrolene sodium and other small molecules targeting the endoplasmic reticulum for the treatment of Wolfram syndrome.

**Trial registration:** ClinicalTrials.gov Identifier NCT02829268

**Key Points:** *Question:* Is dantrolene sodium safe and effective for the treatment of adult and pediatric patients with Wolfram syndrome?

*Findings:* The results of this open-label clinical trial show that dantrolene sodium is well tolerated by patients with Wolfram syndrome. Although the study was small, a select few patients seemed to have improvements in β-cell function, which might correlate with a positive trend in other outcome measures, including visual acuity and neurological functions.

*Meaning:* Dantrolene sodium is well tolerated by patients with Wolfram syndrome. Some patients may experience an increase in β cell function when taking dantrolene.

*Importance:* Wolfram syndrome is a rare endoplasmic reticulum disorder characterized by insulin-dependent diabetes mellitus, optic nerve atrophy, and progressive neurodegeneration. Although there is currently no treatment to delay, halt, or reverse the progression of Wolfram syndrome, preclinical studies in cell and rodent models suggest that targeting endoplasmic reticulum calcium homeostasis, including dantrolene sodium, is an emerging therapeutic strategy.

*Objective:* The primary objective of the study was to assess the safety and tolerability of dantrolene sodium in adult and pediatric subjects with Wolfram syndrome. Secondary objectives were to evaluate the efficacy of dantrolene sodium on residual pancreatic β-cell functions, visual acuity, quality of life measures related to vision, and neurological functions.

*Design:* Open-label phase 1b/2a trial of dantrolene sodium over a 6-month treatment period.

*Setting:* Single site, academic medical center.

*Participants:* Adult and pediatric subjects with a genetically confirmed diagnosis of Wolfram syndrome.

*Interventions:* All subjects received increasing doses of dantrolene sodium.

*Main Outcomes and Measures:* The safety and tolerability of dantrolene sodium administered orally at the upper end of therapeutic dose range for 6 months, and the efficacy of dantrolene sodium on residual pancreatic β-cell functions using a mixed-meal tolerance test, visual acuity using LogMar scores, quality of life measures related to vision using Visual Functioning Questionnaire – 25, and neurological functions using the Wolfram Unified Rating Scale (WURS) and standard neurological assessments.

*Results:* The results indicate that dantrolene sodium is well tolerated by subjects with Wolfram syndrome. Although the study was small, a select few subjects seemed to have improvements in β-cell function, which might be correlated with a positive trend in visual acuity.

*Conclusions and Relevance:* This study justifies further investigation into using dantrolene sodium and other small molecules targeting the endoplasmic reticulum for the treatment of Wolfram syndrome.

*Trial Registration:* Registered with clinicaltrials.gov, NCT02829268, (https://clinicaltrials.gov/ct2/show/NCT02829268?term=NCT02829268&draw=2&rank=1)

## Introduction

Wolfram syndrome is an ultra-rare, progressive neurodegenerative disorder characterized by juvenile-onset insulin-requiring diabetes mellitus and optic nerve atrophy ^1,2^. Other clinical manifestations of Wolfram syndrome include diabetes insipidus, deafness, neurogenic bladder, and ataxia. Most individuals with Wolfram syndrome have a shortened lifespan due to severe neurological disabilities caused by brain stem and cerebellar atrophy ^3^. There has yet to be a treatment devised which has been shown to provide a cure or slow the insidious progression of this disease. As a result, patients with Wolfram syndrome are currently only offered therapies aimed at treated each aspect of the disease individually.

Since the discovery of *WFS1* as the causative locus for most cases of Wolfram syndrome, research efforts have sought to understand the underlying etiology of this disorder. Our current understanding is that Wolfram syndrome is a prototype of endoplasmic reticulum (ER) disease in humans ^4^. WFS1 is a multi-pass ER transmembrane protein with an established role in the negative regulation of ER stress and the maintenance of cellular calcium homeostasis ^5,6^. While the molecular details of WFS1 function require further study, it is clear that pancreatic β-cells and neurons are particularly affected by, and perhaps especially sensitive to, disease-causing *WFS1* genetic variants. Indeed, previous reports from our lab identified calcium dyshomeostasis as a key mechanism underlying pancreatic β-cell and neuronal cell death in the context of WFS1 depletion ^7-9^. These pre-clinical studies led to identify dantrolene sodium as a potential therapeutic candidate for restoring ER calcium homeostasis and mitigating the progression of Wolfram syndrome.

Dantrolene sodium is a hydantoin derivative skeletal muscle relaxant whose mechanism of action revolves around the inhibition of ryanodine receptors (RyR) on the ER ^10-12^. Although the mechanism of action of dantrolene remains unclear, its effects are well-documented. Dantrolene inhibits ER calcium efflux through RyR, thereby reducing cytosolic calcium and preserving ER calcium ^13^. The primary indication for dantrolene is in the treatment of malignant hyperthermia, which can be an adverse reaction to general anesthesia and is FDA approved for use in both adults and children. Dantrolene has also been used off-label for the treatments of spasticity disorders and cerebral vasospasm ^14,15^. Interestingly, recent studies have proposed a potential role for dantrolene sodium as a treatment for neurodegenerative disorders such as Huntington’s disease ^16^, spinocerebellar ataxia ^17,18^, and Alzheimer’s disease ^19,20^, where ER calcium may play a pivotal role in disease pathogenesis.

Murine and induced pluripotent stem cell (iPSC) models of Wolfram syndrome were treated with dantrolene sodium to determine the feasibility of a clinical trial^8^. After receiving promising results from these pre-clinical studies, our group put together the first ever clinical trial in subjects with Wolfram syndrome. Our research team was particularly sensitive to the unique challenges of performing a clinical trial for a disease as rare as Wolfram syndrome ^21^. These challenges include the small numbers of subjects available to study and the vast heterogeneity of symptoms exhibited by patients with Wolfram syndrome ^22^. Multiple stakeholders were involved in the design of this clinical trial including I-TRAK (a natural history study of neurodegeneration in early Wolfram syndrome, NCT03951298) and various Wolfram syndrome patient/parent advocacy groups as they would be likely sources for recruitment. After these collaborations, an open-label phase 1b/2a trial design was chosen. The primary endpoint of the study was to assess the safety and tolerability of dantrolene sodium in adult and pediatric subjects with Wolfram syndrome. Secondary objectives were also to assess the effect of dantrolene sodium on residual pancreatic β-cell function, visual acuity, neurological function and quality of life measures.

## Results

### Trial population

A total of 22 subjects (6–32 years old) with a genetically confirmed diagnosis of Wolfram syndrome, were screened for enrollment in this study (Figure 1B). Of this group, 21 qualified for baseline laboratory and quality of life assessments in order to begin the run-in regimen of oral dantrolene parallel to ongoing maintenance medications. Two subjects (11% of qualified population) had to be excluded before the 6-month assessment of study outcome measures on dantrolene treatment due to loss to follow-up or personal reasons. The baseline demographic and clinical characteristics of the 19 subjects that completed the trial are shown in Table 1. Subject-specific *WFS1* mutations and clinical data are summarized in Supplementary Table S1. At enrollment 100% of subjects carried a diagnosis of diabetes mellitus. However, only 63% of pediatric subjects and 100% of adult subjects carried a diagnosis of optic atrophy. This pattern is consistent with the documented natural history of Wolfram syndrome, where juvenile-onset diabetes mellitus typically manifests within the first decade, followed by optic atrophy in the second decade of life ^3,23,24^.

**Table 1.** Demographic and Clinical Characteristics of the Study Subjects.

| **Characteristic** | **Pediatric (n = 8)** | **Adult (n = 11)** |
| --- | --- | --- |
| Median age (range) - yr | 13 (6-17) | 23 (18-32) |
| Female sex -no. (%) | 6 (75) | 8 (73) |
| BMI (range) - kg/m<sup>2</sup> | 20.3 (15.3-29.3) | 27.0 (17.8-48.9) |
| Race - no (%) |  |  |
| Caucasian | 7 (88) | 9 (82) |
| Hispanic or Latino | 1 (13) | 2 (18) |
| Median age (range) - yr |  |  |
| Diabetes mellitus | 5 (3-7) | 6 (5-14) |
| Optic Atrophy | 7 (5-14) | 14 (6-20) |
| Diabetes insipidus | 8 (6-12) | 17 (6-26) |
| Hearing loss | 7 (3-16) | 8 (2-16) |

**Figure 1.**
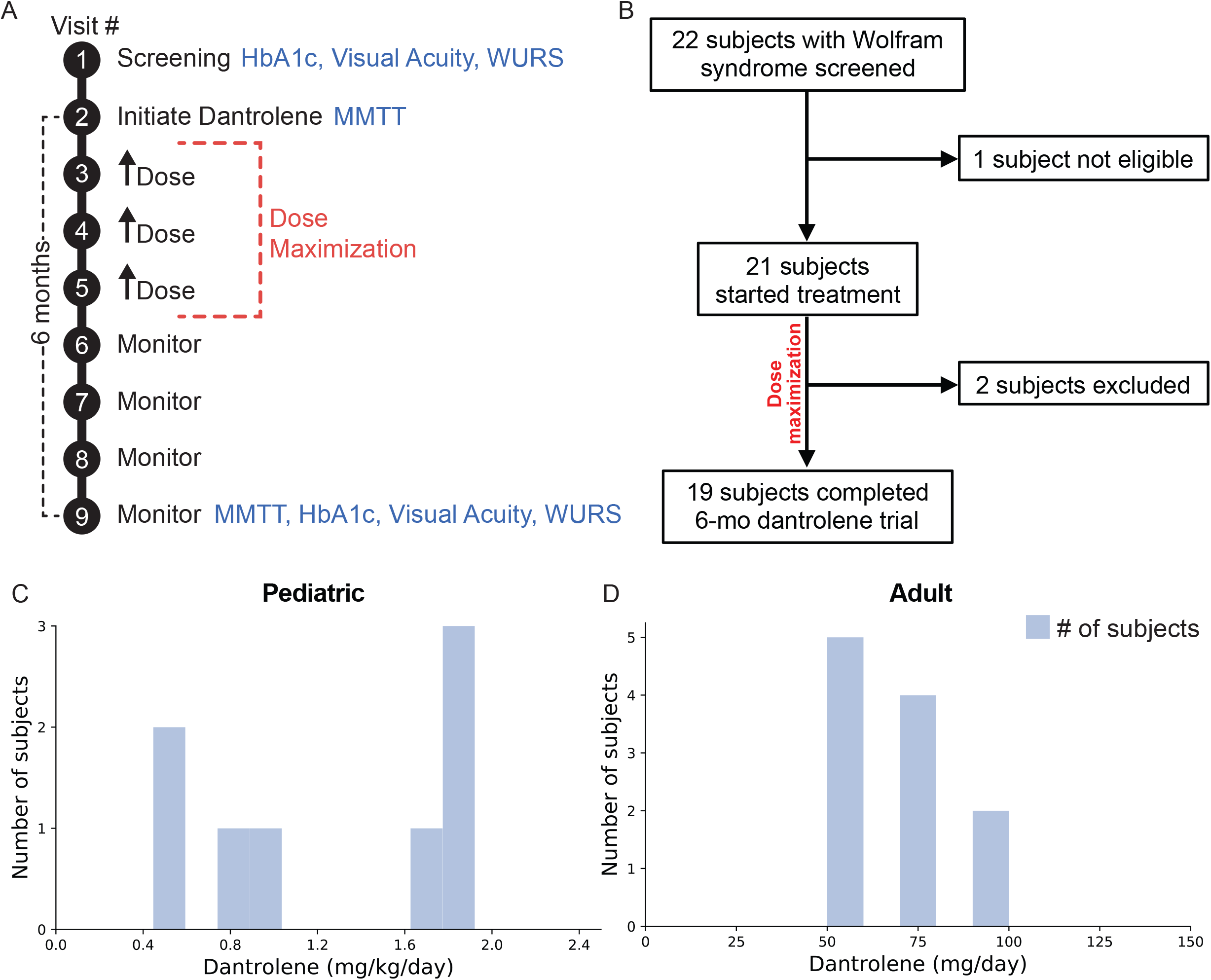
Trial Design, Enrolment, and Retention. A) Schematic of 6-month study. Each study visit is noted by a black circle. Study procedures for secondary endpoints are noted in blue. The dose maximization period for dantrolene sodium is noted by the red dashed lines. B) Enrollment and retention diagram for the subjects enrolled in the study. C) Histogram demonstrating distribution of final tolerated dantrolene doses in pediatric subjects at the end of the study. For pediatric subjects this is expressed as mg/kg/day. D) Histogram demonstrating distribution of final tolerated dantrolene doses in adult subjects at the end of the study. For adult subjects this is expressed as mg/day. For both histograms the blue bars represent numbers of subjects taking a dose, red lines represent the exact doses.

### Safety outcomes

Dantrolene was well-tolerated among pediatric subjects at a final daily dose between 0.5 mg/kg and 2.0 mg/kg, with a maximun daily dose of 100 mg. The mean final daily dose in the pediatric subjects was 1.25 mg/kg/day (Figure 1C). Adults subjects tolerated dantrolene well between 50 mg to 100 mg daily. 5, 4, and 2 subjects tolerated 50, 75, and 100 mg of dantrolene respectively. This resulted in a mean dose of 68.2 mg daily (Figure 1D). These dosing ranges closely approximated therapeutic ranges for dantrolene when used to treat spasticity ^25^.

Adverse events were stratified into three categories based on their likelihood of being attributed to the study drug. These categories included adverse events attributed directly to dantrolene, events known to occur commonly in patients with Wolfram syndrome, and non-specific events not easily attributed to dantrolene or Wolfram syndrome. The most common adverse dantrolene related events observed in pediatric and adult subjects were mild fatigue and diarrhea. The most common Wolfram syndrome related events were mild hypoglycemia, and headaches. These symptoms affected at least 25% of the total study population (Table 2). Hepatoxicity and weakness, the most serious known side effects of dantrolene, were not very prevalent in our study population. Elevated liver enzymes were observed in 2 subjects (11% of total population) and weakness was self-reported by 4 subjects (21% of total population). Quantitative assessments of strength prior to and at each subsequent trial visit after dantrolene administration showed no significant loss in grip strength during 6-months of dantrolene treatment (Supplementary Figure 1). No clinically significant changes in laboratory measures or in findings from physical examinations were noted during enrollment in this study. No significant EKG changes were observed in subjects during the run-in period or thereafter. Additionally, no subject discontinued the trial regimen due to adverse effects.

**Table 2.** Adverse Events During the Study.

| **Adverse Event: no. (%)** | **Pediatric** | **Adult** |
| --- | --- | --- |
| Dantrolene related |  |  |
| Fatigue | 3 (38) | 5 (45) |
| Diarrhea | 2 (25) | 3 (27) |
| Weakness | 3 (38) | 1 (9) |
| Dizziness | 2 (25) | 1 (9) |
| Elevated Hepatic Enzymes | 0 (0) | 2 (18) |
| Nausea | 0 (0) | 2 (18) |
| GI upset | 1 (13) | 1 (9) |
| Rash | 1 (13) | 1 (9) |
| Wolfram related |  |  |
| Hypoglycemia (mild) | 3 (38) | 4 (36) |
| Headaches | 2 (25) | 4 (36) |
| Hyponatremia | 1 (13) | 3 (27) |
| Hyperglycemia | 2 (25) | 1 (9) |
| Urinary tract infection | 1 (13) | 3 (27) |
| Hyperkalemia | 1 (13) | 0 (0) |
| Urinary retention | 1 (13) | 0 (0) |
| Non-specific |  |  |
| Influenza | 1 (13) | 1 (9) |
| Rhinovirus infection | 1 (13) | 0 (0) |
| Knee infection | 1 (13) | 0 (0) |
| Tics | 1 (13) | 0 (0) |
| Knee effusions | 0 (0) | 1 (9) |
| Hit by car | 0 (0) | 1 (9) |
| Pneumonia | 0 (0) | 0 (0) |
| Otitis media | 0 (0) | 0 (0) |

### Secondary outcomes

#### -Markers of β-cell function

To assess the effect of dantrolene on glycemia and remaining β-cell function, HbA1c and 30-minute mixed-meal stimulated C-peptide were monitored at baseline and after 6-months of dantrolene treatment (Figure 1A). Mean HbA1c across all subjects remained stable between dantrolene initiation and after 6-months of treatment (7.4 ± 0.2 %, *p*-value 0.63). Subgroup analyses of adult and pediatric subjects also demonstrated no significant change in HbA1c (7.4 ± 0.2 %) (Figure 2A). Mean fasting C-peptide levels of the total study cohort also remained stable during this period (0.27 ± 0.07 ng/mL at 6-months of treatment compared to 0.27 ± 0.06 ng/mL at baseline, *p*-value 0.95). At the conclusion of the study, mean stimulated C-peptide levels were not significantly higher compared to the pre-treatment baseline (0.64 ± 0.14 ng/mL after 6-months of treatment compared to 0.52 ± 0.10 ng/mL at baseline, *p*-value 0.14) (Figure 2B). Supplementary Figure 2 demonstrates subject specific change in fasting and stimulated C-peptide over the 6-month study period. When looking at all subjects, ΔC-peptide (the change in C-peptide between 0 and 30 minutes) was not significantly increased. Mean ΔC-peptide was 0.37 ± 0.07 ng/mL after 6-months of treatment, compared to 0.25 ± 0.04 ng/mL at baseline (*p*-value 0.18) (Figure 3A and Supplementary Table S2).

**Figure 2.**
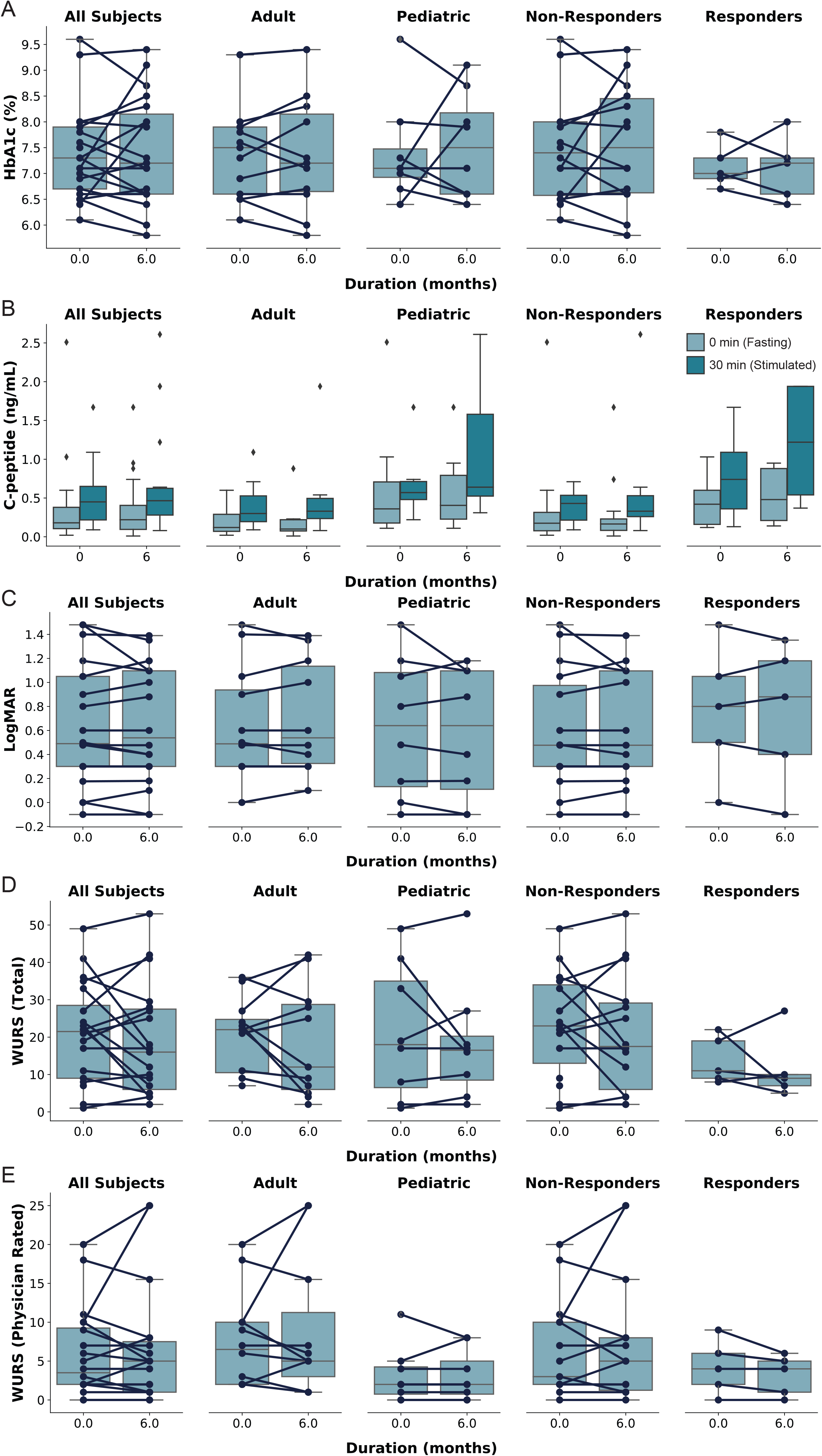
Secondary study endpoints. A) Hemoglobin a1c (HbA1c). B) C-peptide during a mixed meal tolerance text. Light boxes represent fasting results, while dark boxes represent 30-minute (stimulated) values. C) LogMAR (a measure of visual acuity). Lower score correlates to more accurate vision. D) Wolfram Unified Rating Scale (WURS) Score. E) Physician rated subsection of the WURS. Higher WURS scores represent more severe disease. All study subjects are broken down into adult and pediatric subgroups. Responders are differentiated from non-responders by having a change in Δ C-peptide (ΔΔ C-peptide) ≥ 0.1 ng/mL over the study period (see Figure 3).

**Figure 3.**
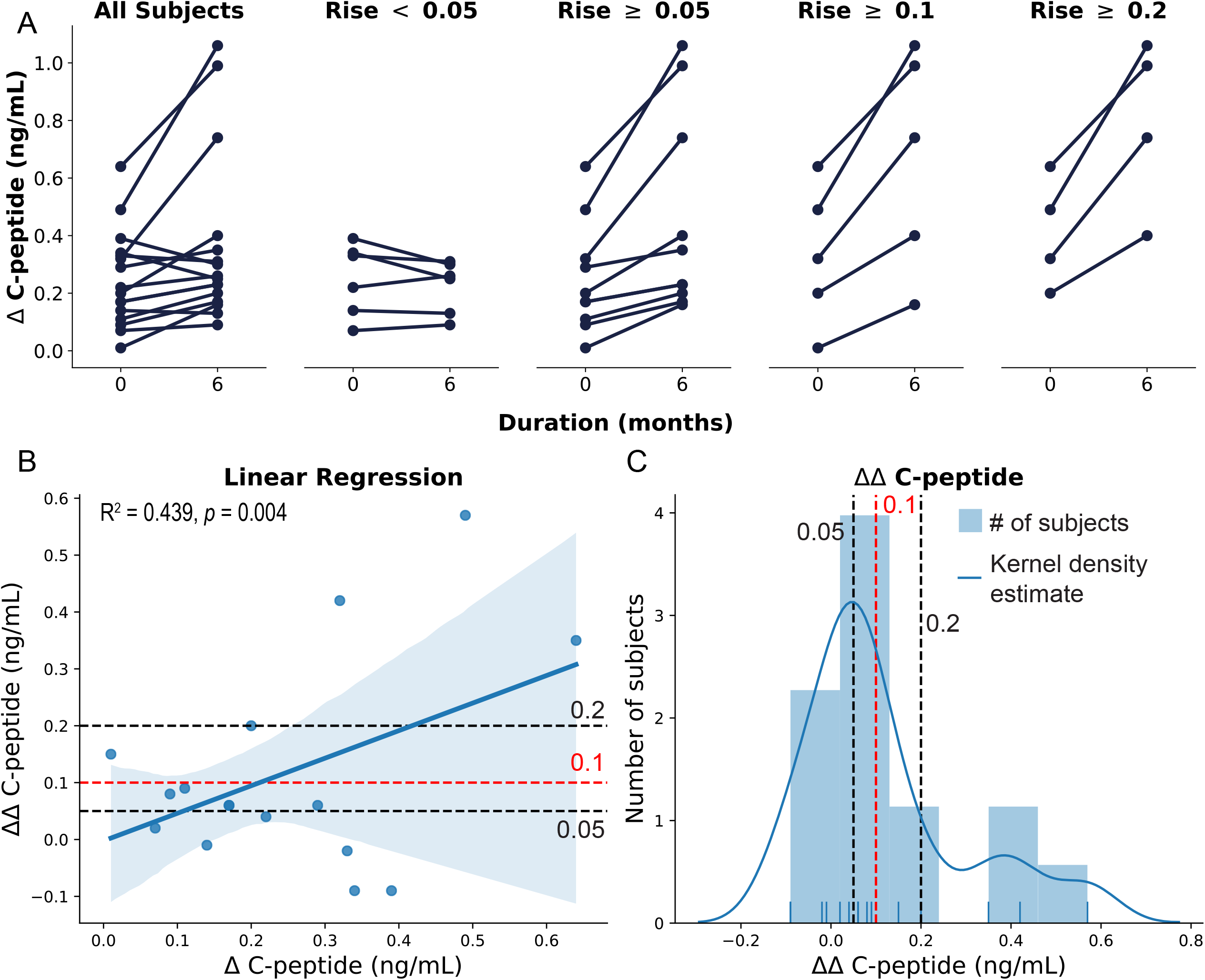
Subgroup analysis to determine responders vs. non-responders. A) Δ C-peptide was plotted between all subjects. Then the change in Δ C-peptide (ΔΔ C-peptide) was calculated for each subject over the course of the study. Subjects were stratified based on a ΔΔ C-peptide < 0.05, ≥ 0.05, ≥ 0.1, and ≥ 0.2 ng/mL respectively. B) Linear regression analysis demonstrates a significant positive relationship between baseline Δ C-peptide and ΔΔ C-peptide. C) Histogram demonstrating the distribution of ΔΔ C-peptide in the study population. Solid blue line demonstrates the kernel density estimate. The different cutoff values for ΔΔ C-peptide (0.05, 0.1, and 0.2 ng/mL) are plotted on both (C) and (D). Based on these analyses a ΔΔ C-peptide ≥ 0.1 ng/mL (depicted in red) was chosen, and these subjects are defined as ‘Responders’ throughout the remainder of the analysis.

Additional markers of β-cell function were performed including measuring proinsulin along with C-peptide during the mixed meal tolerance testing. Insulinogenic index ^26^ and area under the curve (AUC) C-peptide / AUC Glucose ^27^ were calculated for each study subject. No significant differences were found in any of these categories (Supplementary Table S2).

#### -Subgroup analyses

Additional subgroup analysis was performed in the study population to determine if there was subset of subjects who had the most beneficial response to dantrolene. Our hypothesis was that subjects who possessed the greatest degree of β cell function at baseline, would have the greatest glycemic benefit from dantrolene. Therefore, we examined the change in ΔC-peptide (ΔΔC-peptide) over the course of the study to approximate changes in β cell responsiveness. ΔΔC-peptide was calculated by subtracting the ΔC-peptide at baseline from the ΔC-peptide at 6-months. In order to test our hypothesis, we divided the subjects based on increasing cutoffs of ΔΔC-peptide (0.05, 0.1, and 0.2 ng/mL). We noted that 8 subjects had a ΔΔC-peptide ≥ 0.05 ng/mL, 5 subjects had a ΔΔC-peptide ≥ 0.1 ng/mL, and 4 subjects had a ΔΔC-peptide ≥ 0.2 ng/mL. The remaining 6 subjects had a ΔΔC-peptide <0.05 ng/mL or were missing data necessary for calculation of ΔΔC-peptide (Figure 3A). Review of these data suggested that our hypothesis was correct, as subjects with higher ΔC-peptide to begin with, tended to have a higher ΔC-peptide at 6 months and higher slope (ΔΔC-peptide). To further test this relationship, we performed linear regression analysis demonstrating a statistically significant (R^2^ 0.439, *p*-value 0.004) positive relationship between baseline ΔC-peptide, and (ΔΔC-peptide) (Figure 3B). A histogram was created to visualize the distribution of ΔΔC-peptide amongst the study subjects (Figure 3C). The 0.05, 0.1, and 0.2 ng/mL cutoffs were superimposed upon this the linear regression and histograms. Based on previous literature ^28,29^, we decided that the cutoff of a ΔΔC-peptide ≥ 0.1 ng/mL is likely to be of clinical significance, for subjects with Wolfram syndrome. This fit the overall distribution of data, as 5 subjects (subject ID #4, 8, 10, 17, and 22), met this criterion. They represented 26.3% of the total study population. These subjects included 3 adult and 2 pediatric individuals. The ΔΔC-peptide ≥ 0.1 ng/mL cutoff is henceforth used to classify subjects as responders versus non-responders.

As our hypothesis is that subjects with increased β cell function had a more robust response to dantrolene, we set out to determine if any baseline measurement had predictive value. This could help clinicians, caring for patients with Wolfram, determine if dantrolene could be beneficial. Therefore basal, stimulated, and ΔC-peptide data collected from subjects prior to starting dantrolene were analyzed to determine which of these criteria could best be used. Histograms and receiver-operator-characteristic (ROC) curves were created based on these data. A positive control using ΔΔC-peptide which, by design, demonstrated 100% sensitivity and specificity using a ΔΔC-peptide value ≥ 0.12 ng/mL was also created (Supplementary Figure 3D,H). Baseline measurements of β-cell function had a modest predictive value. Unstimulated (Basal) C-peptide had the highest area under the curve (AUC), demonstrating a peak of 60% sensitivity, and 100% specificity when using a cutoff of 0.38 ng/mL. Similarly, stimulated C-peptide demonstrated 60% sensitivity and 100% specificity when using a cutoff of 0.725 ng/mL. ΔC-peptide had the lowest AUC, demonstrating 40% sensitivity and 100% specificity when using a cutoff of 0.44 ng/mL (Supplementary Figure 3A-C, E-G; Supplementary Table S3)

There were no significant differences between responders and non-responders in terms of HbA1c at the beginning (7.1 ± 0.2 % and 7.5 ± 0.2 % respectively, *p*-value 0.49) of the study and after 6-months of treatment (7.1 ± 0.3 % and 7.6 ± 0.3 % respectively, *p-*value 0.42). Prior to treatment with dantrolene, responder subjects had higher fasting C-peptide compared to non-responders (0.47 ± 0.17 ng/dL compared to 0.18 ± 0.03 ng/dL, *p-*value 0.03). By design, responder subjects demonstrated statistically significant increases in fasting C-peptide (0.53 ± 0.17 ng/dL to 0.15 ± 0.006 ng/dL), stimulated C-peptide (1.20 ± 0.33 ng/dL to 0.38 ± 0.05 ng/dL *p-*value 0.003), and ΔC-peptide (0.67 ± 0.17 ng/dL to 0.23 ± 0.02 ng/dL *p-*value 0.002) after the 6-month treatment period. Similar patterns were seen in between responders and non-responders when measuring proinsulin, insulinogenic index, and AUC C-peptide / AUC Glucose. C-peptide to glucose ratio seemed to be elevated in the pediatric and responder groups compared to adults and non-responders respectively (Supplementary Figure 4, Supplementary Table S4).

#### -Markers of visual acuity

To evaluate the effect of dantrolene treatment on visual acuity and vision-related quality of life, participants underwent ophthalmologic examination at screening, and at baseline, and after 6 months of dantrolene treatment. No significant changes in visual acuity were observed across subjects or age groups as a function of dantrolene treatment (Figure 2C, Supplementary Table S2). Of note, subject 12 had a LogMAR = 3 throughout the study period, which equates to functional blindness. As a result, subject 12’s data was excluded from analysis. Supplementary Figure 5 demonstrates the LogMAR data including subject 12. Subgroup analyses did not identify any significant differences between adult and pediatric subjects or subjects deemed to be non-responders or responders. Correspondingly, subjects reported no significant improvements in vision-related quality of life as measured by the NEIVFQ-25 (Supplementary Table S5).

Linear regression analysis was performed comparing ΔΔ C-peptide to LogMAR. This resulted in a slight, but statistically significant negative correlation (R^2^ 0.136, *p*-value 0.023) (Supplementary Figure 6A).

#### -Disease severity and quality of life

Overall disease severity was assessed in subjects prior to, and 6-months after starting dantrolene treatment via WURS assessment ^30^. There were no differences in total WURS disease severity, or mean physician rated physical exam scores across the 6 months of the study. Subgroup analysis noted that pediatric subjects demonstrated statistically insignificant lower physician rated WURS (3.1 ± 1.3 at baseline and 3.1 ± 1.2 after 6-months) (*p*-value 1.00) than adult subjects (7.6 ± 1.8 at baseline and 8.8 ± 2.7 after 6-months) (*p-*value 0.71) (*p-*value 0.09 and 0.11 at baseline and 6-months respectively when comparing pediatric to adult subjects). Subjects deemed to be responders also demonstrated statistically insignificant lower physician rated WURS scores when comparing baseline (4.2 ± 1.6) and 6-months (3.2 ± 1.2) (*p-*value 0.62). Non-responders saw a statistically insignificant increase in physician rated WURS scores when comparing baseline (6.3 ± 1.6) to 6-months of treatment (7.5 ± 2.3) (*p-*value 0.67) (*p-*value 0.48 and 0.28 at baseline and 6-months respectively when comparing non-responder to responder subjects) (Figure 2, and Supplementary Tables S2 and S4). Similarly, pediatric subjects displayed no significant changes in physical or psychosocial health domains as measured by the PedsQL questionnaire between screening and 6 months of dantrolene therapy (Supplementary Table S6) and adult subjects did not show differences in physical or mental health metrics when assessed by the SF-36v (Supplementary Table S7).

Similar to the LogMAR data, linear regression analysis was performed correlating ΔΔ C-peptide to Total WURS and physician rated WURS. These regression plots appeared to demonstrate a negative correlation, but did not reach statistical significance (R2 0.085, *p*-value 0.117 and R2 0.021, *p-*value 0.444 for total WURS and physician rated WURS respectively) (Supplementary Figure 6 B-C).

## Discussion

In this study, we evaluate the safety and tolerability of dantrolene sodium as a therapeutic approach for Wolfram syndrome. Our preclinical studies show that dantrolene improves β-cell and neuronal cell survival in mouse and patient iPSC models of this disease^8^. To translate these findings to humans, we conducted the first clinical trial in pediatric and adult subjects with Wolfram syndrome in a 6-month study of dantrolene sodium (NCT02829268). We identified a tolerable range of oral dantrolene dosing of 0.5mg/kg/day to 2mg/kg/day for pediatric subjects and 50mg/day to 100mg/day for adults. Overall, dantrolene was very well tolerated, and aside from mild fatigue and diarrhea, no clinically significant adverse events were reported.

Admittedly, this proof of concept study ran into many of the same issues that plague early clinical trials for rare diseases ^21^. As the incidence of Wolfram syndrome is so rare, it is difficult to recruit a large enough sample size in order to detect a statistically significant difference in secondary measures of β-cell function, visual acuity, or quality of life. In order to aid with recruitment, our study team collaborated with existing natural history studies and patient/parent organizations. Particularly, patient/parent organizations expressed a strong desire for a potential therapeutic option, as there are currently no approved treatments aimed at slowing the progression of Wolfram syndrome. Through the design of the study, potential study subjects lobbied strongly against a blinded or placebo-controlled study design. This helped inform our decision toward an open-label phase 1b/2a design, as a positive outcome would clear a path for more widespread adoption of a drug aimed at slowing the progression of Wolfram syndrome that is at least proven to be safe, if not necessarily effective in all individuals. The final challenge facing this study is the vast clinical heterogeneity seen in the spectrum of individuals with Wolfram syndrome ^31^. Independent of age, some individuals are more severely affected or progress more rapidly than others. Our hypothesis is that these differences may be based on the severity of the *WFS1* gene variants. For example, individuals with missense mutations, may have a less severe course compared to large deletions or non-sense mutations. This clinically and genetically heterogeneous population makes it more challenging to infer cause and effect relationships when studying a potential drug. As a result, our strategy has been to target the underlying cellular defect (ER calcium depletion) that is unified amongst all patients with Wolfram syndrome ^32^.

With the above challenges in mind, there remain many shortcomings of this study. As it is an uncontrolled study, certain parameters measured are susceptible to confounding by the placebo effect. Due to the small sample sizes there were no statistically significant differences in β-cell function or disease severity. For these reasons, this study does not posit that dantrolene improves β-cell function or disease severity. Instead, it identifies safe doses for treatment of adult and pediatric subjects with Wolfram syndrome, highlights the side effect profile of dantrolene in this population and argues that further investigation of dantrolene, or investigational agents with a similar mechanism of action, are warranted in a randomized, double-blind, placebo-controlled study. With this caveat in mind, this study also suggests that dantrolene requires further investigation in the context of β-cell function and neurodegeneration.

Perhaps the most salient question arising from this study is whether dantrolene improves human β-cell function in Wolfram syndrome. Mean stimulated C-peptide was not significantly different when looking at all subjects. However, parsing subjects by age reveals that pediatric subjects had a statistically insignificant increase in C-peptide. Subgroup analyses, suggests that subjects with better baseline β-cell function (earlier on in the progression of the disease and β-cell loss) may have the most benefit from dantrolene sodium (Figure 3B). Interestingly, linear regression analysis also demonstrated that subjects with the greatest increase in β-cell function tended to have improved visual acuity and less severe disease (Supplementary Figure 6A). These data suggest that the greatest beneficiaries of dantrolene treatment may be newly diagnosed pediatric subjects who retain a significant degree of β-cell function. The ROC analysis (Supplementary Figure 3) suggests that having a fasting C-peptide > 0.38 ng/mL, can help predict if a subject will have an greater than 0.1 ng/mL increase Δ C-peptide over 6 months of dantrolene treatment. While a controlled human study is required to assess dantrolene’s efficacy at improving β-cell function, pediatric subjects in our study started to exhibit a trend towards higher stimulated C-peptide levels after 6-months of sustained dantrolene treatment. Adult subjects, in contrast, show a negligible increase in mean stimulated C-peptide levels throughout their duration of dantrolene treatment. These data suggest that dantrolene may be more effective in pediatric subjects, possibly because these subjects have a larger surviving subpopulation of functional β-cells during this initial phase of their disease process. Evidently, adult subjects also secrete very low levels of insulin, but dantrolene did not seem to significantly enhance β-cell function in this group.

The significance of these small elevations in C-peptide is quite interesting when comparing Wolfram syndrome to type 1 diabetes. Recently, there has been a growing body of literature suggesting that there is clinical benefit from a very small degree of residual β-cell function. Oram and colleagues published a population-based study in the United Kingdom, demonstrating that 8% of subjects had a urinary C-peptide-to-creatinine ratio ≥ 0.2 nmol/mmol ^29^. This study and another follow-up study in 2019 demonstrated that this persistent micro-secretion of C-peptide is associated with fewer complications of diabetes and less hypoglycemia ^28^. Contrasting these populations, subjects with Wolfram syndrome tend to have much higher C-peptide compared to type 1 diabetes. Notably the preserved C-peptide group in the 2019 study had a mean stimulated C-peptide of 114 pmol/L (0.3443 ng/mL). This is compared to a mean stimulated C-peptide of 0.52 ng/mL (205 pmol/L) in our study population with Wolfram syndrome. These data suggest that small, statistically insignificant, increases in C-peptide may be clinically significant. Anecdotally, some of the study investigators noticed that the subject’s insulin needs decreased during the study, but this was not systematically evaluated in this study. Additionally, many subjects wearing a continuous glucose monitors noticed more stable glycemic patterns. As a result, we suggest that future studies of dantrolene or similar agents track changes in total daily insulin dose (with percentage basal versus bolus) and analyze continuous glucose monitor tracings (*i*.*e*. time-in-range, time-in-hypoglycemia, and standard deviation).

Similar to β-cell function, over 6 months of treatment with dantrolene sodium there were no significant differences in markers of visual acuity. Markers of disease severity including WURS score and other pediatric and adult quality of life measures did not significantly change over 6 months of treatment with dantrolene sodium. However, we noted that pediatric subjects tended to have lower physician rated WURS scores compared to their adult counterparts.

We postulate that efficacy of dantrolene may be linked to the nature of the *WFS1* mutations in the individual subjects. Over time, and with further experience with dantrolene, perhaps dantrolene can be part of a personalized medicine approach for patients with Wolfram syndrome.

In summary, this study suggests that dantrolene sodium is safely tolerated by subjects with Wolfram syndrome. Although the study was small, a select few subjects seemed to have improvements in β-cell function. Therefore, this study justifies further investigation into using dantrolene sodium and other ER-calcium stabilizers for the treatment of Wolfram syndrome.

## Methods

### Study approval

Subjects, and their parent or legal guardian, as appropriate, provided written, informed consent before participating in this study, which was approved by the Human Research Protection Office at Washington University School of Medicine in St. Louis, MO (IRB ID #201607006).

### Trial participants

Subjects who met all of the following criteria were eligible for enrolment:

1. A definitive diagnosis of Wolfram syndrome, as determined by the following:
  a. Documented functionally relevant recessive mutations on both alleles of the *WFS1* gene or,
  b. A dominant mutation on one allele of the *WFS1* gene based on historical test results (if available) or from a qualified laboratory at screening.
2. The subject is at least 5 years of age (biological age) at the time of written informed consent.
3. The subject, subject’s parent(s), or legally authorized guardian(s) must have voluntarily signed an Institutional Review Board/Independent Ethics Committee-approved informed consent form after all relevant aspects of the study have been explained and discussed with the subject. The guardians’ consent and subject’s assent, as relevant, must be obtained.

### Study protocol

Dantrolene sodium was dispensed to the study subjects via the Washington University’s clinical trials pharmacy. Subjects were instructed to take the dantrolene by mouth. Subjects enrolled in this study underwent a run-in period for dose maximization (Figure 1A). Adult subjects were started on up to 25 mg dantrolene daily for seven days, then doubled in dose on a weekly basis up to a maximum of 200mg dantrolene daily. Pediatric subjects (< 18-years old) were started on up to 0.5 mg/kg dantrolene daily (maximum 25 mg) for seven days, then doubled in dose on a weekly basis up to a maximum of 2mg/kg dantrolene daily (maximum 200 mg), with no dose change if weight fell within ± 3% of the original dosing weight. Dosing calendars were maintained by the study subjects to ensure adherence to the study drug.

### Safety assessment and outcomes measures

Baseline screening procedures included complete physical exam, standard clinical laboratory tests (serum chemistry, liver function tests, hematology, and urinalysis), and 12-lead ECG. Subjects underwent formal visual acuity testing by the co-authors of the study who are either optometrists or ophthalmologists. At baseline, each subject underwent the Wolfram Unified Rating Scale (WURS) ^30^. The 30-minute mixed meal tolerance test was performed to assess base-line β cell functions as described before ^24^. The mixed meal consisted of 6 ml/kg (maximum 360 ml) of Boost® (Nestle) consumed over a maximum of 5 minutes. After the overnight fasting, blood for glucose and C-peptide measurement was drawn at time 0 (fasting) and 30 minutes after the Boost. If a subject’s fasting glucose exceeded 250 mg/dL, the test was not performed, but fasting glucose and C-peptide were obtained. An ECG was performed before and 4-hours after the first dose of dantrolene was administered during the run-in period, then again at 2-months and 6-months. Best-corrected visual acuity was assessed by Snellen optotype and converted to LogMar score ^33^. Vision-related quality of life was assessed in all subjects at screening and after 6 months of dantrolene by the National Eye Institute’s 25-item Visual Function Questionnaire (VFQ-25) ^34^. Functional activities of daily living were assessed in pediatric subjects by the Pediatric Quality of Life Inventory (PedsQL) (https://www.pedsql.org/) ^35^, while the SF-36v2 (https://www.optum.com/) was used to measure self-reported functional health and well-being of adults at baseline and after 6-months of dantrolene therapy ^36^. If no safety concerns were identified at screening, subjects began the 3-week dose maximization period of dantrolene sodium. All baseline screening procedures were repeated again at 6-months of treatment to ensure subject safety and assess dantrolene tolerability. Grip strength was measured at each visit bilaterally using a digital hand dynamometer (CAMRY). A final safety follow-up visit was conducted at 28 days (+/- 7 days) after the last outcome measure evaluation in order to collect additional information on adverse events, concomitant medications, therapies and procedures. For subjects who discontinued the study prior to the first outcomes measures evaluation, safety follow-up visit was conducted within 28 days (+/- 7 days) after the last administration of dantrolene sodium.

### Statistics

Secondary outcome measures (HbA1c, Glucose, C-peptide, Proinsulin, Insulinogenic Index, AUC C-peptide/ AUC Glucose, LogMAR, and WURS) were reported with the mean result ± the standard error of the mean (SEM). T-tests were performed on these secondary outcome measures when comparing the same group using scipy (https://www.scipy.org) and pandas (https://pandas.pydata.org/index.html) programming libraries. Paired t-tests were used when comparing the same group (all-subjects, adults, pediatrics, non-responders, responders) at 0 and 6 months. Linear regression analyses (including R^2^ and *p-*values) were constructed using the ordinary least squares method using the statsmodels(https://www.statsmodels.org/devel/about.html#about-statsmodels) programming library. Subjects who dropped out of the study or who did not complete a secondary outcome measure were excluded from the analysis. Independent t-tests were performed when comparing adult to pediatric subjects and non-responders to responder subjects. A *p-*value < 0.05 was considered significant for all analyses. Figures were constructed with matplotlib (https://matplotlib.org) and seaborn (https://seaborn.pydata.org) programming libraries or GraphPad Prism 8 software (https://www.graphpad.com/scientific-software/prism/). Receiver-operator-characteristic (ROC) curves and calculations were constructed using GraphPad Prism 8 software.

## Data Availability

All the data are included in the main figures and tables and supplementary information.

## Author contributions

FU designed the study. BAM, NHW, TP, RB, and TH advised on the design of the study. ANS recruited participants and ANS and SH managed the study. TP, RB, JH, LT, SIS, BAM, NHW, and FU examined subjects. SH, CMB, and KK collected the data. CM, DA, SIS, TH, and FU analyzed the data. HG and TH advised on the statistical analysis. DA and SIS wrote the first draft of the manuscript and all the authors revised it critically and approved the final version. DA and SIS are co-first authors.

## Acknowledgements

This work was partly supported by the grants from the National Institutes of Health (NIH)/NIDDK (DK112921, DK113487, DK020579), NIH/ National Center for Advancing Translational Sciences (NCATS) (TR002065, TR000448) and philanthropic supports from the Silberman Fund, the Ellie White Foundation for the Rare Genetic Disorders, the Snow Foundation, the Unravel Wolfram Syndrome Fund, the Stowe Fund, the Eye Hope Foundation, and the Feiock Fund to F. Urano. Research reported in this publication was also supported by the Washington University Institute of Clinical and Translational Sciences grant UL1TR002345 from the NIH/NCATS. The content is solely the responsibility of the authors and does not necessarily represent the official view of the NIH. The authors thank all the members of the Washington University Wolfram Syndrome Study and Research Clinic for their support (https://wolframsyndrome.dom.wustl.edu) and all the participants in the Wolfram syndrome International Registry and Clinical Study, Research Clinic, and Clinical Trials for their time and efforts. D. Abreu was supported by the NIH training grant (F30DK111070).

## Supplementary Information

**Supplementary Figure 1. Grip strength of each study subject during the study**. A) Right hand B) Left hand.

**Supplementary Figure 2. C-peptide data from each study subject over the course of the study**. Blue represents baseline (fasting) C-peptide. Orange represents 30-minute (stimulated) C-peptide. C-peptide is in ng/mL.

**Supplementary Figure 3. Predictive value of baseline data**. Histograms plotting A) Basal (fasting) C-peptide, B) Stimulated (30-minute) C-peptide, C) Δ C-peptide, and D) ΔΔ C-peptide (as a positive control). Receiver-Operator-Characteristic (ROC) curves were created from these data in order to plot the sensitivity vs. specificity of being a responder based on E) Basal (fasting) C-peptide, F) Stimulated (30-minute) C-peptide, G) Δ C-peptide, and H) ΔΔ C-peptide (as a positive control). Area under the curve (AUC) was calculated for each ROC curve. A red dashed line indicates the point with the highest sensitivity and specificity within each ROC curve. This cutoff value is illustrated by a vertical line and annotated on the corresponding histogram.

**Supplementary Figure 4. Additional markers of β-cell function**. A) Proinsulin collected during a mixed meal tolerance text. B) Insulinogenic Index. C) Area Under the Curve (AUC) C-peptide / AUC Glucose. D) C-peptide to Glucose Ratio All study subjects are broken down into adult and pediatric subgroups. Light boxes represent fasting results, while dark boxes represent 30-minute (stimulated) values. Responders are differentiated from non-responders by having a change in Δ C-peptide (ΔΔ C-peptide) ≥ 0.1 ng/mL over the course of the study (see Supplementary Figure 3).

**Supplementary Figure 5. LogMAR visual acuity plot including subject 12**. Subject 12 was excluded from the analysis as they are blind with a LogMar = 3.

**Supplementary Figure 6. Linear regression analysis**. Linear regression analysis comparing ΔΔ C-peptide to (A) LogMAR visual acuity, (B) total WURS, and (C) physician rated WURS. R^2^ and p-values are demonstrated in the top right corner of each panel.

**Supplementary Table S1**. **Genetic and Clinical Characteristics of the Study Subjects**.

**Supplementary Table S2. Secondary Study Endpoints**.

**Supplementary Table S3. Sensitivity and Specificity**

Table based on different cutoff values for Basal C-peptide, Stimulated C-peptide, Δ C-peptide, and ΔΔ C-peptide (positive control). Corresponds to Supplementary Figure 3.

**Supplementary Table S4. Table comparing subgroup analyses at each timepoint**.

**Supplementary Table S5. Vision-related quality of life by the NEIVFQ-25**.

**Supplementary Table S6. Pediatric Quality of Life (PedsQL) questionnaire**.

**Supplementary Table S7. Physical and mental health metrics as assessed by the SF-36v**.

