## Supplementary Information for "A Phase 1b/2a Clinical Trial of Dantrolene Sodium in Patients with Wolfram Syndrome"

**Supplementary Figure 1. Grip strength of each study subject during the study.** A) Right hand B) Left hand.

**Supplementary Figure 2. C-peptide data from each study subject over the course of the study.** Blue represents baseline (fasting) C-peptide. Orange represents 30-minute (stimulated) C-peptide. C-peptide is in ng/mL.

**Supplementary Figure 3. Predictive value of baseline data.** Histograms plotting A) Basal (fasting) C-peptide, B) Stimulated (30-minute) C-peptide, C) Δ C-peptide, and D) ΔΔ C-peptide (as a positive control). Receiver-Operator-Characteristic (ROC) curves were created from these data in order to plot the sensitivity vs. specificity of being a responder based on E) Basal (fasting) C-peptide, F) Stimulated (30-minute) C-peptide, G) Δ C-peptide, and H) ΔΔ C-peptide (as a positive control). Area under the curve (AUC) was calculated for each ROC curve. A red dashed line indicates the point with the highest sensitivity and specificity within each ROC curve. This cutoff value is illustrated by a vertical line and annotated on the corresponding histogram.

**Supplementary Figure 4. Additional markers of β-cell function.** A) Proinsulin collected during a mixed meal tolerance text. B) Insulinogenic Index. C) Area Under the Curve (AUC) C-peptide / AUC Glucose. D) C-peptide to Glucose Ratio All study subjects are broken down into adult and pediatric subgroups. Light boxes represent fasting results, while dark boxes represent 30-minute (stimulated) values. Responders are differentiated from non-responders by having a change in Δ C-peptide (ΔΔ C-peptide) $\geq 0.1$ ng/mL over the course of the study (see Supplementary Figure 3).

**Supplementary Figure 5. LogMAR visual acuity plot including subject 12.** Subject 12 was excluded from the analysis as they are blind with a LogMar = 3.

**Supplementary Figure 6. Linear regression analysis.** Linear regression analysis comparing ΔΔ C-peptide to (A) LogMAR visual acuity, (B) total WURS, and (C) physician rated WURS. $R^2$ and p-values are demonstrated in the top right corner of each panel.

**Supplementary Table S1 . Genetic and Clinical Characteristics of the Study Subjects.**

**Supplementary Table S2. Secondary Study Endpoints.**

**Supplementary Table S3. Sensitivity and Specificity**

Table based on different cutoff values for Basal C-peptide, Stimulated C-peptide, Δ C-peptide, and ΔΔ C-peptide (positive control). Corresponds to Supplementary Figure 3.

**Supplementary Table S4. Table comparing subgroup analyses at each timepoint.**

**Supplementary Table S5. Vision-related quality of life by the NEIVFQ-25.**

**Supplementary Table S6. Pediatric Quality of Life (PedsQL) questionnaire.**

**Supplementary Table S7. Physical and mental health metrics as assessed by the SF-36v.**

Supplementary Figure 1

A

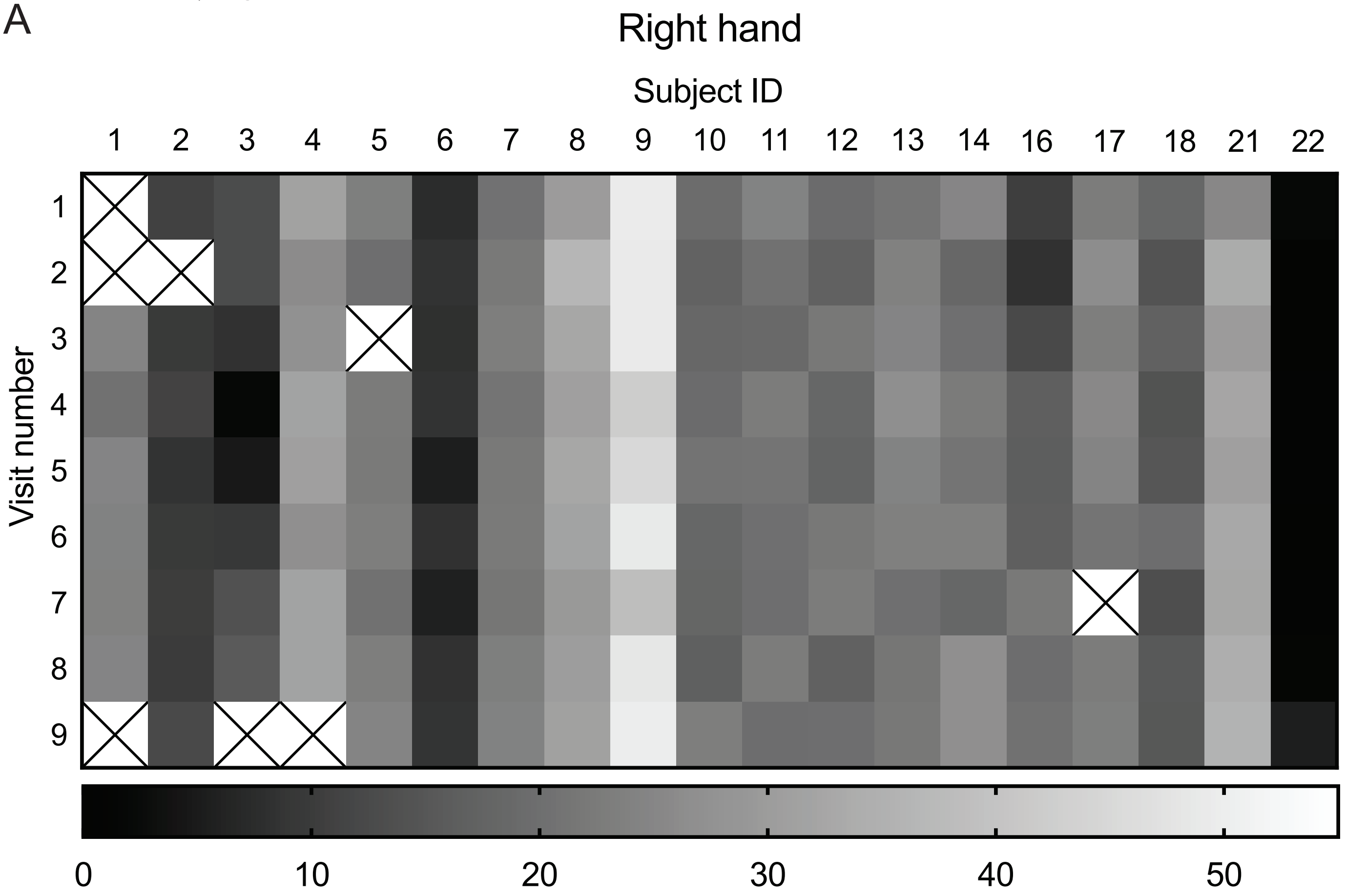


B

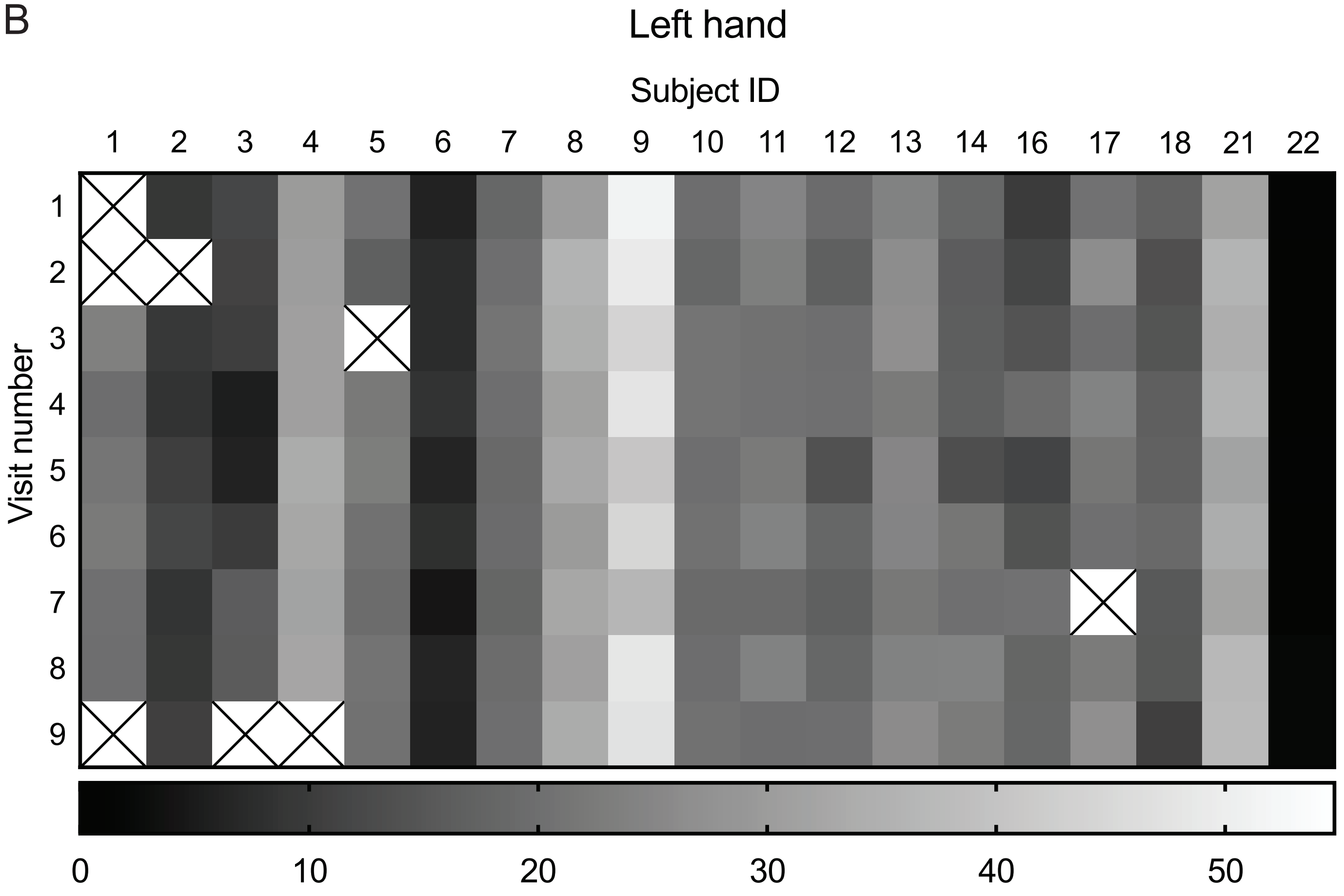

Supplementary Figure 2

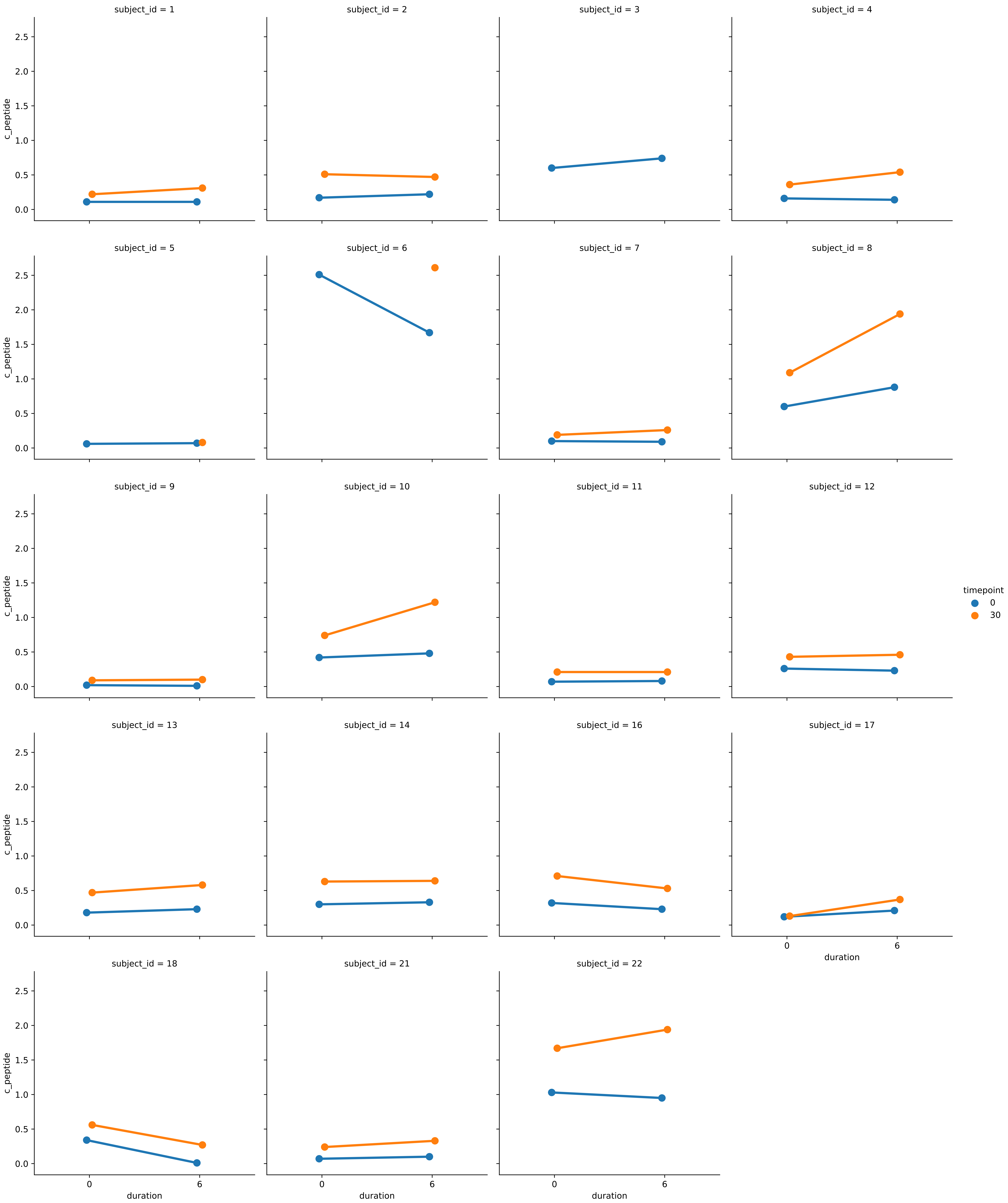

Supplementary Figure 3

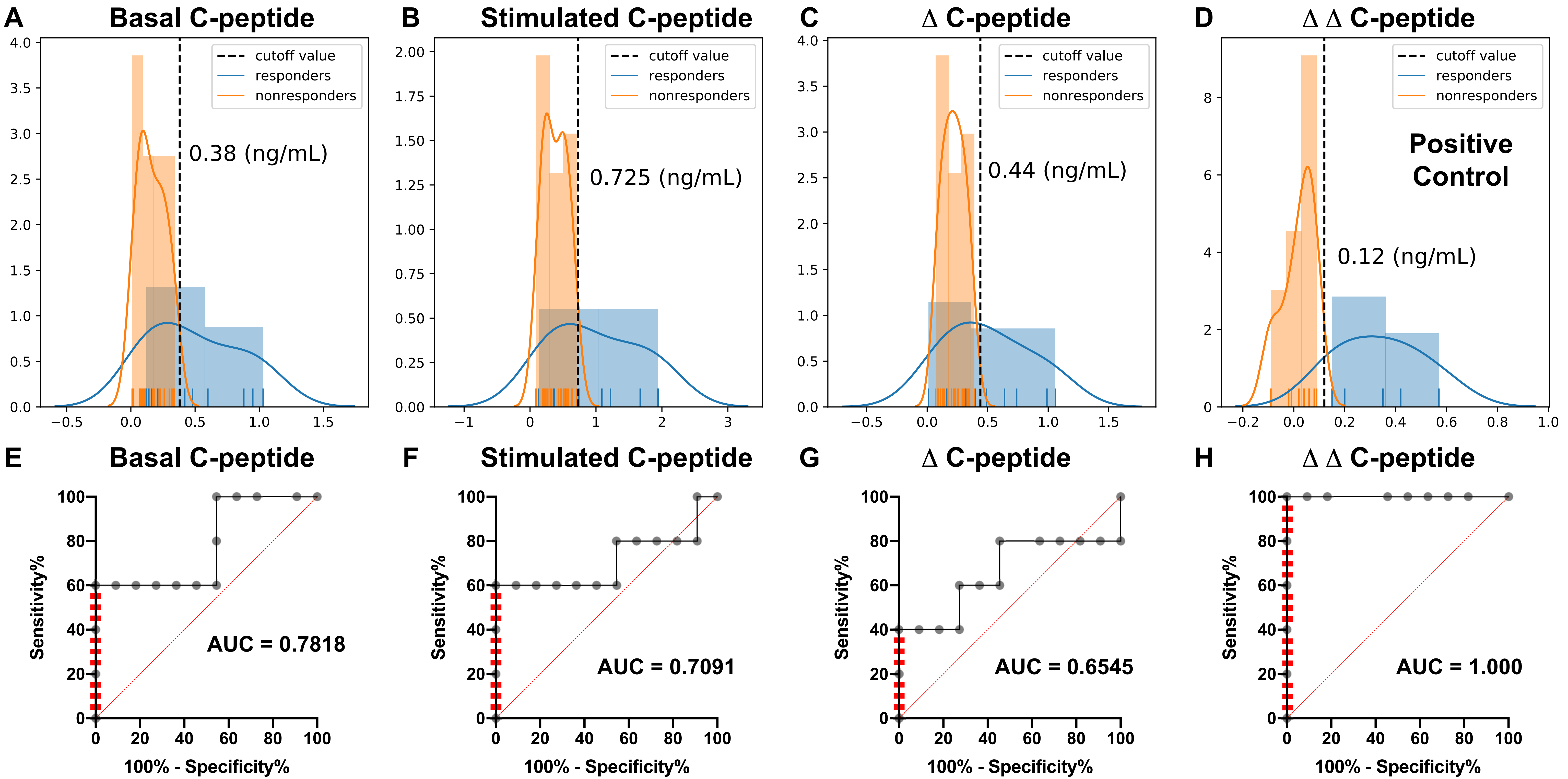

Supplementary Figure 4

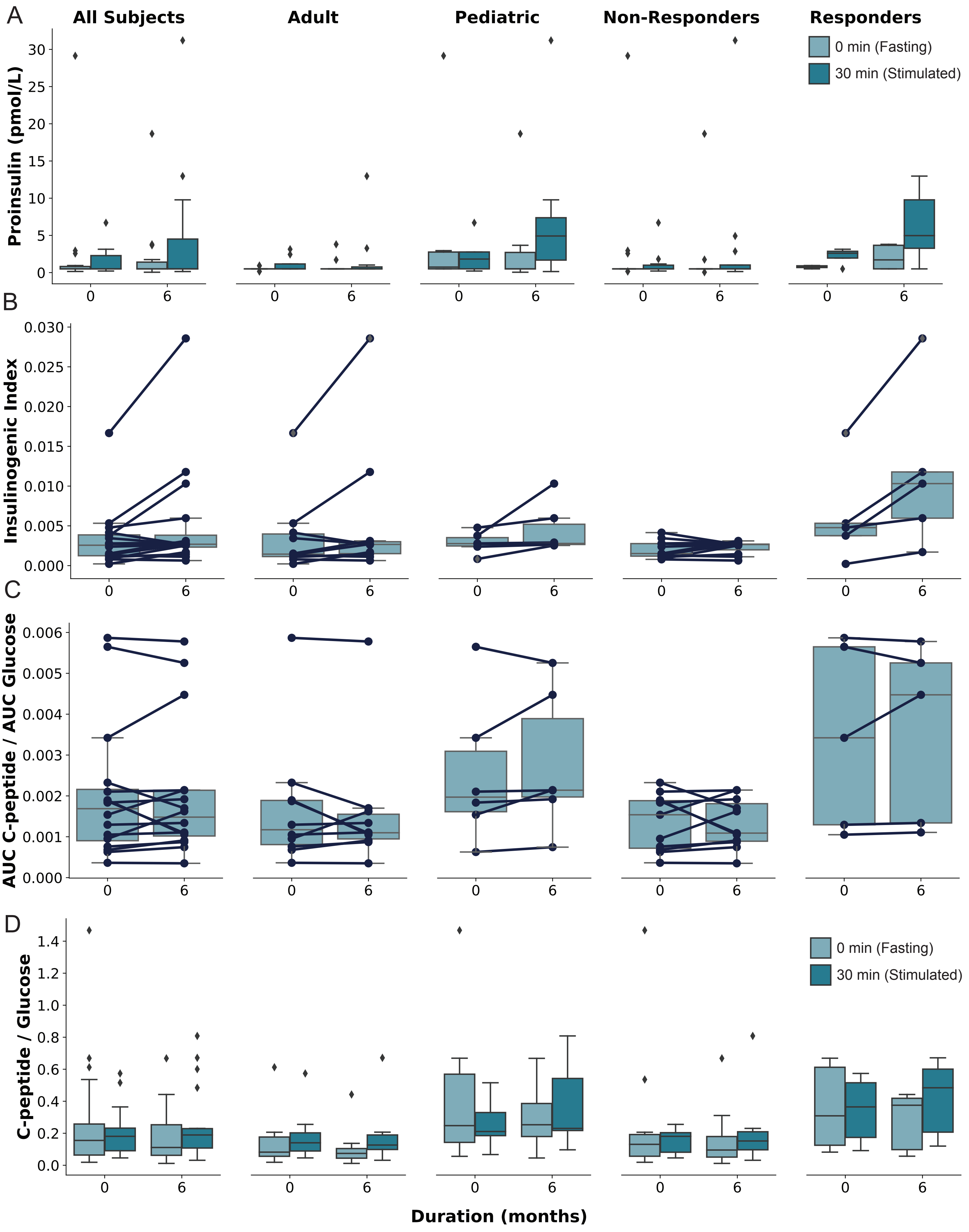

Supplementary Figure 5

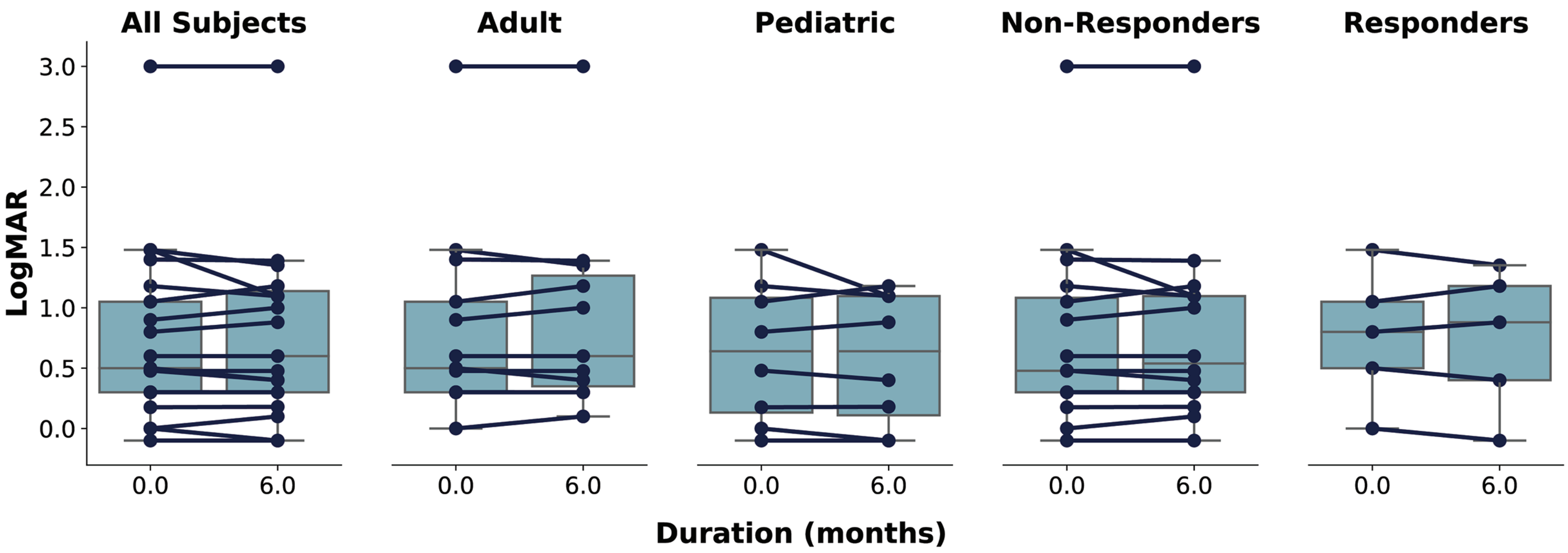

Supplementary Figure 6

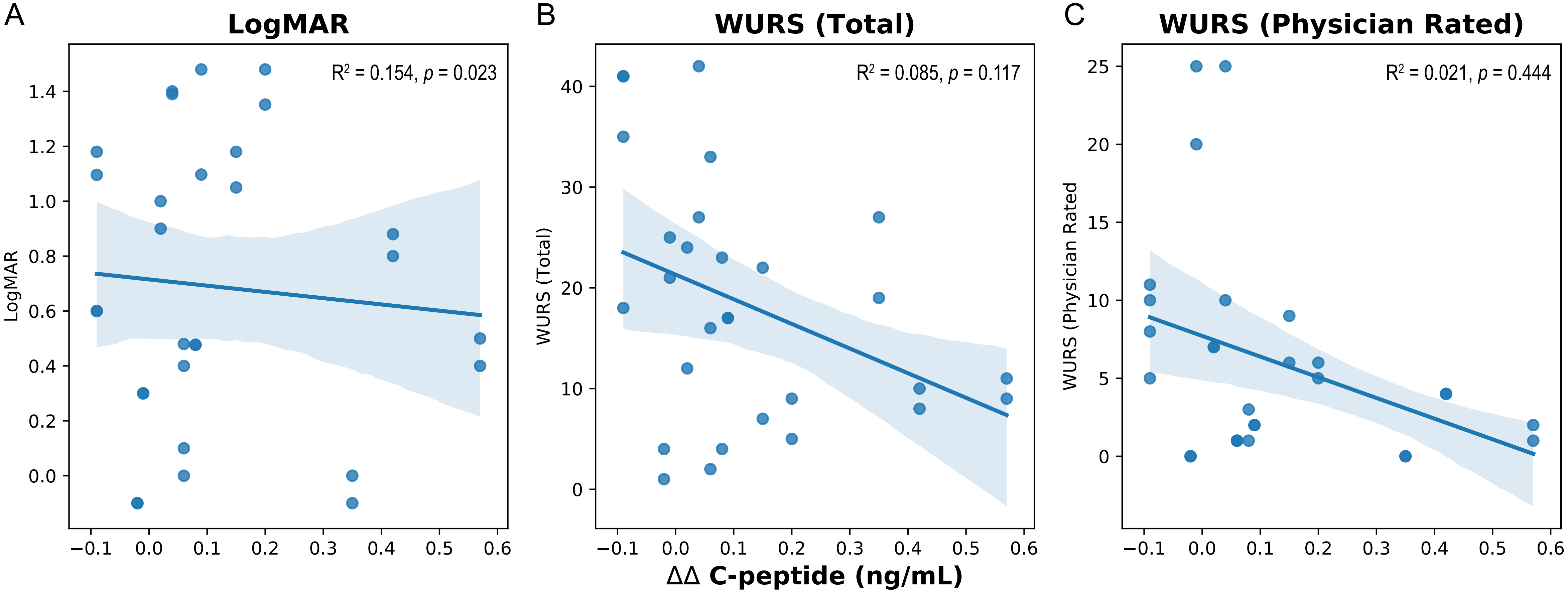

| | WFS1 | | Age at Diagnosis | | | |
|---|---|---|---|---|---|---|
| Subject ID | Allele 1 | Allele 2 | Diabetes Mellitus | Optic Atrophy | Diabetes Insipidus | Hearing Loss |
| 1 | p.(R147Gfs*17) | p.(W540*) | 3 | 7 | 12 | 9 |
| 2 | p.(L200Rfs*87) | p.(E752*) | 5 | 5 | 7 | 7 |
| 3 | p.(E169K) | p.(E169K) | 5 | 6 | 6 | 6 |
| 4 | p.(V412Sfs*29) | p.(V415del) | 5 | 6 | 6 | 6 |
| 5 | p.(L200P) | p.(R232P) | 5 | 15 | -- | 16 |
| 6 | p.(L200P) | p.(R232P) | 7 | -- | -- | 3 |
| 7 | p.(K253*) | p.(F883Sfs*68) | 7 | 20 | 22 | -- |
| 8 | p.(E202G) | p.(D211N) | 14 | 13 | -- | -- |
| 9 | p.(G107E) | p.(R629W) | 6 | 17 | 17 | 7 |
| 10 | p.(F883Sfs*68) | -- | 6 | 9 | 10 | 16 |
| 11 | p.(H313Y) | -- | 4 | 13 | -- | 2 |
| 12 | p.(E273*) | p.(W613*) | 6 | 8 | 11 | 9 |
| 13 | p.(E202G) | p.(F350del) | 7 | 14 | -- | -- |
| 14 | p.(E202G) | p.(F350del) | 5 | -- | -- | -- |
| 16 | p.(L282P) | p.(W613*) | 6 | 18 | 17 | 16 |
| 17 | p.(S790*) | p.(L567_F568del) | 7 | 12 | -- | -- |
| 18 | p.(E169K) | p.(V415del) | 9 | 15 | ? | 8 |
| 21 | p.(F413_V415del) | p.(F883Sfs*68) | 5 | 14 | 16 | -- |
| 22 | p.(R42Efs*101) | p.(F883Sfs*68) | 4 | -- | -- | -- |

**Supplementary Table S1. Genetic and Clinical Characteristics of the Study Subjects**

| Class | All Subjects | | | | | Adults | | | | | Pediatrics | | | | | Non-Responders | | | | | Responders | | | | |
|---|---|---|---|---|---|---|---|---|---|---|---|---|---|---|---|---|---|---|---|---|---|---|---|---|---|
| Duration (Months) | 0 | SEM | 6 | SEM | *p*-value | 0 | SEM | 6 | SEM | *p*-value | 0 | SEM | 6 | SEM | *p*-value | 0 | SEM | 6 | SEM | *p*-value | 0 | SEM | 6 | SEM | *p*-value |
| HbA1c (%) | 7.4 | 0.2 | 7.4 | 0.2 | 0.88 | 7.4 | 0.2 | 7.4 | 0.3 | 0.94 | 7.4 | 0.4 | 7.6 | 0.4 | 0.77 | 7.5 | 0.2 | 7.6 | 0.3 | 0.82 | 7.1 | 0.2 | 7.1 | 0.3 | 0.91 |
| Fasting Glucose (mg/dL) | 145.5 | 9.4 | 165.3 | 16.2 | 0.30 | 140.4 | 13.6 | 161.5 | 23.4 | 0.45 | 154.0 | 11.1 | 171.5 | 21.2 | 0.48 | 149.9 | 11.1 | 148.0 | 20.1 | 0.93 | 135.8 | 18.5 | 203.2 | 20.4 | 0.04 |
| Stimulated Glucose (mg/dL) | 243.3 | 12.5 | 263.8 | 13.3 | 0.27 | 223.6 | 12.9 | 255.9 | 18.4 | 0.17 | 276.2 | 20.0 | 277.0 | 18.3 | 0.98 | 257.0 | 10.8 | 253.4 | 17.8 | 0.86 | 213.2 | 30.0 | 286.8 | 13.6 | 0.06 |
| Δ Glucose (mg/dL) | 97.8 | 9.7 | 98.6 | 7.5 | 0.95 | 83.2 | 11.3 | 94.4 | 10.9 | 0.48 | 122.2 | 13.5 | 105.5 | 8.8 | 0.32 | 107.1 | 7.0 | 105.4 | 6.7 | 0.86 | 77.4 | 26.6 | 83.6 | 18.4 | 0.85 |
| Fasting C-peptide (ng/mL) | 0.27 | 0.06 | 0.27 | 0.07 | 0.98 | 0.21 | 0.06 | 0.20 | 0.08 | 0.94 | 0.37 | 0.14 | 0.39 | 0.12 | 0.92 | 0.18 | 0.03 | 0.15 | 0.03 | 0.56 | 0.47 | 0.17 | 0.53 | 0.17 | 0.79 |
| Stimulated C-peptide (ng/mL) | 0.52 | 0.10 | 0.64 | 0.14 | 0.50 | 0.40 | 0.10 | 0.50 | 0.17 | 0.61 | 0.71 | 0.21 | 0.86 | 0.25 | 0.65 | 0.39 | 0.06 | 0.38 | 0.05 | 0.91 | 0.80 | 0.27 | 1.20 | 0.33 | 0.38 |
| Δ C-peptide (ng/mL) | 0.25 | 0.04 | 0.37 | 0.07 | 0.18 | 0.19 | 0.05 | 0.30 | 0.09 | 0.29 | 0.34 | 0.07 | 0.47 | 0.13 | 0.38 | 0.21 | 0.03 | 0.23 | 0.02 | 0.66 | 0.33 | 0.11 | 0.67 | 0.17 | 0.14 |
| Fasting Proinsulin (pmol/L) | 0.8 | 0.2 | 1.0 | 0.3 | 0.60 | 0.6 | 0.1 | 1.0 | 0.3 | 0.24 | 1.0 | 0.5 | 1.2 | 0.6 | 0.16 | 0.7 | 0.2 | 0.6 | 0.1 | 0.21 | 0.8 | 0.1 | 2.0 | 0.7 | 0.31 |
| Stimulated Proinsulin (pmol/L) | 1.6 | 0.5 | 2.7 | 0.9 | 0.31 | 1.1 | 0.3 | 2.1 | 1.2 | 0.31 | 2.4 | 1.2 | 3.9 | 1.5 | 0.83 | 1.3 | 0.6 | 1.1 | 0.4 | 0.86 | 2.2 | 0.6 | 6.3 | 2.2 | 0.25 |
| Δ Proinsulin (pmol/L) | 0.8 | 0.3 | 1.7 | 0.7 | 0.24 | 0.5 | 0.3 | 1.1 | 0.9 | 0.36 | 1.4 | 0.7 | 2.7 | 1.1 | 0.55 | 0.5 | 0.4 | 0.6 | 0.4 | 0.87 | 1.4 | 0.5 | 4.3 | 1.6 | 0.25 |
| Insulinogenic Index | 0.003 | 0.001 | 0.005 | 0.002 | 0.112 | 0.004 | 0.002 | 0.006 | 0.003 | 0.179 | 0.003 | 0.001 | 0.005 | 0.001 | 0.134 | 0.002 | 0.000 | 0.002 | 0.000 | 0.669 | 0.006 | 0.003 | 0.012 | 0.005 | 0.128 |
| AUC C-peptide / AUC Glucose | 0.002 | 0.000 | 0.002 | 0.000 | 0.994 | 0.002 | 0.001 | 0.002 | 0.000 | 0.165 | 0.003 | 0.001 | 0.003 | 0.001 | 0.128 | 0.001 | 0.000 | 0.001 | 0.000 | 0.492 | 0.003 | 0.001 | 0.004 | 0.001 | 0.388 |
| BMI ($kg/m^2$) | 24.5 | 1.7 | 24.2 | 1.9 | 0.94 | 27.1 | 2.3 | 27.0 | 2.7 | 0.79 | 20.3 | 1.5 | 20.3 | 1.6 | 0.89 | 25.4 | 2.0 | 25.2 | 2.4 | 0.61 | 21.5 | 2.3 | 21.3 | 2.4 | 0.70 |
| LogMAR | 0.7 | 0.2 | 0.8 | 0.2 | 0.90 | 0.8 | 0.2 | 0.9 | 0.2 | 0.76 | 0.6 | 0.2 | 0.6 | 0.2 | 0.85 | 0.7 | 0.2 | 0.8 | 0.2 | 0.87 | 0.8 | 0.2 | 0.7 | 0.3 | 0.95 |
| WURS | 20.8 | 3.0 | 18.5 | 3.4 | 0.61 | 20.5 | 2.8 | 18.6 | 4.5 | 0.72 | 21.3 | 6.4 | 18.4 | 5.7 | 0.74 | 23.1 | 3.7 | 21.0 | 4.3 | 0.70 | 13.8 | 2.8 | 11.6 | 3.9 | 0.66 |
| WURS (Physician Rated) | 5.8 | 1.3 | 6.4 | 1.7 | 0.78 | 7.6 | 1.8 | 8.8 | 2.7 | 0.71 | 3.1 | 1.3 | 3.1 | 1.2 | 1.00 | 6.3 | 1.6 | 7.5 | 2.3 | 0.67 | 4.2 | 1.6 | 3.2 | 1.2 | 0.62 |

**Supplementary Table S2. Secondary study endpoints**

| Basal C-peptide | Sensitivity% | Specificity% | Likelihood ratio | Stimulated C-peptide | Sensitivity% | Specificity% | Likelihood ratio | Δ C-peptide | Sensitivity% | Specificity% | Likelihood ratio | ΔΔ C-peptide | Sensitivity% | Specificity% | Likelihood ratio |
|---|---|---|---|---|---|---|---|---|---|---|---|---|---|---|---|
| > 0.04500 | 100 | 9.091 | 1.1 | > 0.1100 | 100 | 9.091 | 1.1 | > 0.04000 | 80 | 0 | 0.8 | > -0.05500 | 100 | 18.18 | 1.222 |
| > 0.08500 | 100 | 27.27 | 1.375 | > 0.1600 | 80 | 9.091 | 0.88 | > 0.08000 | 80 | 9.091 | 0.88 | > -0.01500 | 100 | 27.27 | 1.375 |
| > 0.1050 | 100 | 36.36 | 1.571 | > 0.2000 | 80 | 18.18 | 0.9778 | > 0.1000 | 80 | 18.18 | 0.9778 | > 0.005000 | 100 | 36.36 | 1.571 |
| > 0.1150 | 100 | 45.45 | 1.833 | > 0.2150 | 80 | 27.27 | 1.1 | > 0.1250 | 80 | 27.27 | 1.1 | > 0.03000 | 100 | 45.45 | 1.833 |
| > 0.1400 | 80 | 45.45 | 1.467 | > 0.2300 | 80 | 36.36 | 1.257 | > 0.1550 | 80 | 36.36 | 1.257 | > 0.05000 | 100 | 54.55 | 2.2 |
| > 0.1650 | 60 | 45.45 | 1.1 | > 0.3000 | 80 | 45.45 | 1.467 | > 0.1850 | 80 | 54.55 | 1.76 | > 0.07000 | 100 | 81.82 | 5.5 |
| > 0.1750 | 60 | 54.55 | 1.32 | > 0.3950 | 60 | 45.45 | 1.1 | > 0.2100 | 60 | 54.55 | 1.32 | > 0.08500 | 100 | 90.91 | 11 |
| > 0.2200 | 60 | 63.64 | 1.65 | > 0.4500 | 60 | 54.55 | 1.32 | > 0.2550 | 60 | 63.64 | 1.65 | > 0.1200 | 100 | 100 | |
| > 0.2800 | 60 | 72.73 | 2.2 | > 0.4900 | 60 | 63.64 | 1.65 | > 0.3050 | 60 | 72.73 | 2.2 | > 0.1750 | 80 | 100 | |
| > 0.3100 | 60 | 81.82 | 3.3 | > 0.5350 | 60 | 72.73 | 2.2 | > 0.3250 | 40 | 72.73 | 1.467 | > 0.2750 | 60 | 100 | |
| > 0.3300 | 60 | 90.91 | 6.6 | > 0.5950 | 60 | 81.82 | 3.3 | > 0.3350 | 40 | 81.82 | 2.2 | > 0.3850 | 40 | 100 | |
| > 0.3800 | 60 | 100 | | > 0.6700 | 60 | 90.91 | 6.6 | > 0.3650 | 40 | 90.91 | 4.4 | > 0.4950 | 20 | 100 | |
| > 0.5100 | 40 | 100 | | > 0.7250 | 60 | 100 | | > 0.4400 | 40 | 100 | | | | | |
| > 0.8150 | 20 | 100 | | > 0.9150 | 40 | 100 | | > 0.5650 | 20 | 100 | | | | | |
| | | | | > 1.380 | 20 | 100 | | | | | | | | | |

**Supplementary Table S3. Sensitivity and Specificity Table (Corresponds to Supplementary Figure 3E-G).**

| Duration (Months) | 0 | | | | | 6 | | | | | 0 | | | | | 6 | | | | |
|---|---|---|---|---|---|---|---|---|---|---|---|---|---|---|---|---|---|---|---|---|
| Class | Adults | SEM | Pediatrics | SEM | *p*-value | Adults | SEM | Pediatrics | SEM | *p*-value | Non-Responders | SEM | Responders | SEM | *p*-value | Non-Responders | SEM | Responders | SEM | *p*-value |
| BMI ($kg/m^2$) | 27.1 | 2.3 | 20.3 | 1.5 | 0.05 | 27.0 | 2.7 | 20.3 | 1.6 | 0.07 | 25.4 | 2.0 | 21.5 | 2.3 | 0.32 | 25.2 | 2.4 | 21.3 | 2.4 | 0.37 |
| HbA1c (%) | 7.4 | 0.2 | 7.4 | 0.4 | 0.97 | 7.4 | 0.3 | 7.6 | 0.4 | 0.70 | 7.5 | 0.2 | 7.1 | 0.2 | 0.49 | 7.6 | 0.3 | 7.1 | 0.3 | 0.42 |
| Fasting Glucose (mg/dL) | 140.4 | 13.6 | 154.0 | 11.1 | 0.50 | 161.5 | 23.4 | 171.5 | 21.2 | 0.78 | 149.9 | 11.1 | 135.8 | 18.5 | 0.51 | 148.0 | 20.1 | 203.2 | 20.4 | 0.12 |
| Stimulated Glucose (mg/dL) | 223.6 | 12.9 | 276.2 | 20.0 | 0.04 | 255.9 | 18.4 | 277.0 | 18.3 | 0.46 | 257.0 | 10.8 | 213.2 | 30.0 | 0.11 | 253.4 | 17.8 | 286.8 | 13.6 | 0.26 |
| Δ Glucose (mg/dL) | 83.20 | 11.29 | 122.17 | 13.46 | 0.05 | 94.40 | 10.86 | 105.50 | 8.81 | 0.49 | 107.09 | 7.01 | 77.40 | 26.61 | 0.16 | 105.36 | 6.74 | 83.60 | 18.41 | 0.19 |
| Fasting C-peptide (ng/dL) | 0.21 | 0.06 | 0.37 | 0.14 | 0.23 | 0.20 | 0.08 | 0.39 | 0.12 | 0.20 | 0.18 | 0.03 | 0.47 | 0.17 | 0.03 | 0.15 | 0.03 | 0.53 | 0.17 | 0.01 |
| Stimulated C-peptide (ng/dL) | 0.40 | 0.10 | 0.71 | 0.21 | 0.15 | 0.50 | 0.17 | 0.86 | 0.25 | 0.23 | 0.39 | 0.06 | 0.80 | 0.27 | 0.06 | 0.38 | 0.05 | 1.20 | 0.33 | 0.00 |
| Δ C-peptide (ng/dL) | 0.2 | 0.0 | 0.3 | 0.1 | 0.10 | 0.3 | 0.1 | 0.5 | 0.1 | 0.28 | 0.2 | 0.0 | 0.3 | 0.1 | 0.19 | 0.2 | 0.0 | 0.7 | 0.2 | 0.00 |
| Fasting Proinsulin (pmol/L) | 0.6 | 0.1 | 1.0 | 0.5 | 0.24 | 1.0 | 0.3 | 1.2 | 0.6 | 0.73 | 0.7 | 0.2 | 0.8 | 0.1 | 0.95 | 0.6 | 0.1 | 2.0 | 0.7 | 0.01 |
| Stimulated Proinsulin (pmol/L) | 1.1 | 0.3 | 2.4 | 1.2 | 0.19 | 2.1 | 1.2 | 3.9 | 1.5 | 0.38 | 1.3 | 0.6 | 2.2 | 0.6 | 0.40 | 1.1 | 0.4 | 6.3 | 2.2 | 0.01 |
| Δ Proinsulin (pmol/L) | 0.5 | 0.3 | 1.4 | 0.7 | 0.20 | 1.1 | 0.9 | 2.7 | 1.1 | 0.29 | 0.5 | 0.4 | 1.4 | 0.5 | 0.22 | 0.6 | 0.4 | 4.3 | 1.6 | 0.01 |
| Insulinogenic Index | 0.004 | 0.002 | 0.003 | 0.001 | 0.737 | 0.006 | 0.003 | 0.005 | 0.001 | 0.765 | 0.002 | 0.000 | 0.006 | 0.003 | 0.044 | 0.002 | 0.000 | 0.012 | 0.005 | 0.007 |
| AUC C-peptide / AUC Glucose | 0.002 | 0.001 | 0.003 | 0.001 | 0.352 | 0.002 | 0.000 | 0.003 | 0.001 | 0.168 | 0.001 | 0.000 | 0.003 | 0.001 | 0.013 | 0.001 | 0.000 | 0.004 | 0.001 | 0.006 |
| Logmar Score | 0.63 | 0.14 | 0.63 | 0.21 | 1.00 | 0.14 | 0.71 | 0.58 | 0.19 | 0.60 | 0.59 | 0.13 | 0.77 | 0.25 | 0.51 | 0.62 | 0.13 | 0.74 | 0.27 | 0.65 |
| WURS | 20.50 | 2.82 | 21.25 | 6.38 | 0.91 | 18.59 | 4.51 | 18.38 | 5.71 | 0.98 | 23.13 | 3.68 | 13.80 | 2.82 | 0.18 | 20.96 | 4.34 | 11.60 | 3.94 | 0.24 |
| WURS (Physician Rated) | 7.6 | 1.8 | 3.1 | 1.3 | 0.09 | 8.8 | 2.7 | 3.1 | 1.2 | 0.11 | 6.3 | 1.6 | 4.2 | 1.6 | 0.48 | 7.5 | 2.3 | 3.2 | 1.2 | 0.28 |

**Supplementary Table S4. Table comparing subgroup analyses at each timepoint**

| | Duration of Dantrolene Treatment (months) | |
|---|---|---|
| | **0** | **6** |
| **VFQ-25 Domain** | (n=19) | (n=19) |
| General health | 60.5 (4.4) | 56.6 (4.6) |
| General vision | 61.1 (6.4) | 56.8 (5.4) |
| Ocular pain | 81.6 (4.4) | 82.2 (4.2) |
| Near activities | 59.2 (4.7) | 57.0 (4.9) |
| Distance Activities | 65.9 (4.3) | 59.6 (4.5) |
| Vision specific | | |
| Social functioning | 77.6 (5.0) | 77.0 (5.1) |
| Mental health | 67.8 (3.9) | 68.8 (4.1) |
| Role difficulties | 66.4 (5.8) | 73.7 (4.9) |
| Dependency | 63.2 (4.8) | 62.3 (4.7) |
| Driving | 45.5 (13.8) | 52.8 (14.7) |
| Color vision | 68.1 (7.8) | 59.2 (8.6) |
| Peripheral vision | 73.7 (6.8) | 63.2 (7.7) |
| **Composite** | **68.8 (3.1)** | **70.4 (3.2)** |

**Supplementary Table S5. Vision-related quality of life by the NEIVFQ-25**

| | Duration of Dantrolene Treatment (months) | |
|---|---|---|
| | **0** | **6** |
| **Scale - Mean (SEM)** | (n=8) | (n=8) |
| Physical health | 77.7 (6.8) | 80.4 (4.9) |
| Psychosocial health | 65.6 (9.7) | 63.3 (8.1) |
| **Total score** | **69.8 (8.6)** | **69.3 (6.6)** |

**Supplementary Table S6. Pediatric Quality of Life (PedsQL) questionnaire.**

SF-36v Score

| Duration (months) | 0 | 6 | *p*-value |
|---|---|---|---|
| Count | 11 | 12 | |
| Mean (SEM) | 115.4 (2.5) | 108.8 (2.7) | 0.3 |

**Supplementary Table S7. Physical and mental health metrics as assessed by the SF-36v.**
